# Risk of root resorption between Invisalign and fixed orthodontic treatment: A retrospective study

**DOI:** 10.1101/2024.12.05.24318570

**Authors:** Hanadi M. Sabban, Linah Al-Labban, Nouf Baeshen, Khadijah A. Turkistani

**Affiliations:** Division of Oral & Maxillofacial Radiology, Department of Oral Diagnostic Sciences, King Abdulaziz University, Faculty of Dentistry, Jeddah, Saudi Arabia; Private clinics, King Abdulaziz University School of Dental Medicine Graduate, Jeddah, Saudi Arabia; Department of Orthodontics, King Abdulaziz University, Faculty of Dentistry, Jeddah, Saudi Arabia

**Keywords:** Orthodontic treatment, Root resorption, Clear aligners, Invisalign, Fixed appliances, Tooth movement, Panoramic radiographs (OPG), Periapical radiographs, Malocclusion, Orthodontic complications, Retrospective study, Apical root resorption

## Abstract

**Objectives:** The aim of our study is to evaluate and compare the presence and severity of external root resorption in patients treated with either clear aligners (Invisalign) versus conventional fixed orthodontic appliances.

**Material and Methods:** A retrospective study was performed on 203 patients, which includes 30 treated with Invisalign and 173 with fixed appliances. 60 cases (30 per group) were matched based on extraction status, treatment duration, gender, age, and malocclusion classification after inclusion criteria were applied. Radiographic evaluations were conducted with standardized periapical radiographs and Panoramic radiographs (OPG). Comparing radiographs taken before and after treatment allowed for the measurement of the extent of external root resorption.

**Results:** In the matched cases, external root resorption was present in 92% of patients in the Invisalign group and 77% of patients in the fixed appliance group. Maxillary incisors were the most afflicted teeth, particularly in situations involving tooth extractions. The greatest percentage of root resorption (100%) was recorded in Class II malocclusion, which was followed by Class I (84%) and Class III (77%). Although Invisalign treatments demonstrated an increased incidence of root resorption, fixed appliances were associated with more severe root reduction. There were not significant distinctions between the groups, according to statistical analysis (p > 0.05).

**Conclusions:** A frequent adverse effect of orthodontic treatments involving both fixed appliances and Invisalign is external root resorption. Root resorption was more common in Invisalign patients, but it was greater in fixed appliance cases. These results highlight the need for more expansive prospective studies to confirm these findings and enable clinicians minimize side effects associated with treatment.

## 1. Introduction

The aim of this study is to assess the prevalence of root resorption in cases of mild to moderate malocclusion treated with either fixed orthodontic appliances or clear removable aligners (Invisalign). The objective of may help orthodontists choose the best course of treatment for their patients by maintaining effectiveness with reduced side effects. Malocclusion is believed to have an impact on the overall condition of the masticatory system [1]. Improvements in orthodontic materials and techniques have led to the establishment of a variety of treatment options, enabling individualized care that addresses each patient’s needs. Clear aligners, especially Invisalign, have become popular due to their aesthetic appeal, convenience, and patient comfort [2]. These clear, custom-made trays use attachments to improve the accuracy of tooth movements and fit tightly over the teeth. On the other hand, brackets attached to teeth and bonded by archwires that apply constant forces which encourage tooth movement have been referred to as fixed orthodontic appliances. Fixed appliances have been suggested for moderate to severe malocclusions because of their capacity to apply constant forces over prolonged periods of time, even though clear aligners are frequently selected for their hygienic and cosmetic benefits [3].

There are significant biomechanical discrepancies between fixed appliances and clear aligners. Although fixed appliances transmit constant forces, clear aligners produce intermittent forces and can be removed for oral hygiene and consuming food. Furthermore, the way forces are distributed varies: aligners use attachments that distribute forces over multiple points, whereas fixed appliances centralized force transmission [4].

A widely recognized side effect of orthodontic treatment is root resorption, which is defined by the loss of teeth root structure, primarily affecting the cementum and dentin. Irreversible root shortening may result from excessive orthodontic pressures affecting the surrounding bone and periodontal ligament. The level of resorption is greatly determined by variables such the severity of the malocclusion, the amount and duration of applied forces, and the form of each individual root [5].

The effect of treatment intervals on apical root resorption in patients with fixed appliances was investigated by Levander et al. [6]. Their research demonstrated that, in comparison to continuous therapy, a two- to three-month treatment break decreased the amount of root resorption, highlighting the significance of treatment timing to managing root resorption.

For the diagnosis and assessment of root resorption, radiographic imaging is still essential. Cone-beam computed tomography (CBCT), periapical radiographs, and panoramic radiographs are common modalities. Three CBCT devices were examined for radiation exposure levels by Ludlow et al. [7], who also noted differences in exposure according to imaging techniques and device type. Although CBCT provides better diagnostic accuracy, cost and radiation concerns prevent it from being widely used, hence panoramic radiographs are a viable substitute in many orthodontic practices [8].

In orthodontics, classification systems play an essential function in accurate diagnosis and treatment planning. Class I (neutro-occlusion), Class II (disto-occlusion), and Class III (mesio-occlusion) are among the malocclusion classes that are still categorized using Angle’s classification, which was first proposed in 1899 [9]. Space analysis, which examines tooth dimensions and arch length, is also essential for detecting crowding or spacing disorders [10].

## 2. Methods and methods

### 2.1 Ethical Approval and Participants Consent

King Abdulaziz University Dental Hospital’s orthodontic cases’ anonymized medical records were retrospectively analyzed for this study. As part of the institution’s regular clinical protocol, all patients (or their legal guardians for minors) gave written informed consent for treatment and the use of de-identified data for research purposes at the beginning of orthodontic treatment.

The information was retrieved for study purposes between [December 15, 2021, and May 31, 2022]. Prior to analysis, all records were anonymised, and the authors did not have access to any identifying information either during or after the study. The King Abdulaziz University Dental Hospital’s Research Ethics Committee provided ethical approval for this study (Approval number: 161-12-20).

### 2.2 Study Design and Sample Size Calculation

The study compared external root resorption between two orthodontic treatment modalities—fixed appliances and clear aligners (Invisalign)—using a cross-sectional retrospective design. The required number of samples was determined using Cochran’s formula, which guarantees a 95% confidence level with a 5% margin of error. Based on the population estimate of about 3.5 million as published by the General Authority for Statistics in 2021, the first calculation suggested that 385 cases per group would be required for statistical power. The final matched sample addressed sample size restrictions from earlier studies by including 30 cases each group (60 total) to improve statistical reliability.

### 2.3 Data Collection and Patient Selection

The orthodontic treatment cases were selected from King Abdulaziz University Dental Hospital’s archive. Initially, 203 orthodontic cases were found, of which 30 were treated using Invisalign and 173 with fixed equipment. Propensity score matching further narrowed the remaining 71 cases to 60 matched cases (30 per group) after applying inclusion and exclusion criteria.

**Inclusion and Exclusion Criteria**

**Inclusion Criteria:**

- Patients aged 15–60 years at the time of treatment.
- Completed orthodontic treatments with fixed appliances or Invisalign.
- Cases involving mild to moderate malocclusion.
- Treatments completed between 2010 and 2021.
- Availability of both pre-treatment and post-treatment radiographs.

**Exclusion Criteria:**

- Diagnosed bone pathologies.
- Patients younger than 15 or older than 60 at treatment initiation.
- Asymmetrical molar relationships or single-arch treatments.
- Incomplete treatments or ongoing interventions.
- Missing pre-treatment or post-treatment radiographs.
- Use of mini-screws, coils, headgear, or face masks.
- Previous jaw surgeries (e.g., maxillary advancement).
- History of root canal treatments or impacted tooth-related root resorption.

### 2.4 Radiographic Analysis

Digital images stored in King Abdulaziz University dental hospital’s R4 electronic filing system were used for radiography evaluations. Periapical and Panoramic radiographs (OPG) radiographs were obtained during standard clinical visits.

**Peripheral radiographs:** are obtained by a standardized parallel technique that ensures consistent angulation and reduces distortion.

**Panoramic radiographs (OPG):** Despite their known precision limitations, OPGs are used for more comprehensive diagnostic support.

Images taken before and after treatment were compared to determine the amount of root resorption in millimeters, with an emphasis on changes in root length (Figure 1). A calibrated examiner conducted these measurements to guarantee consistency and reliability when assessing resorption across all cases. Images that were unclear or of poor quality were not included.

**Figure 1.**
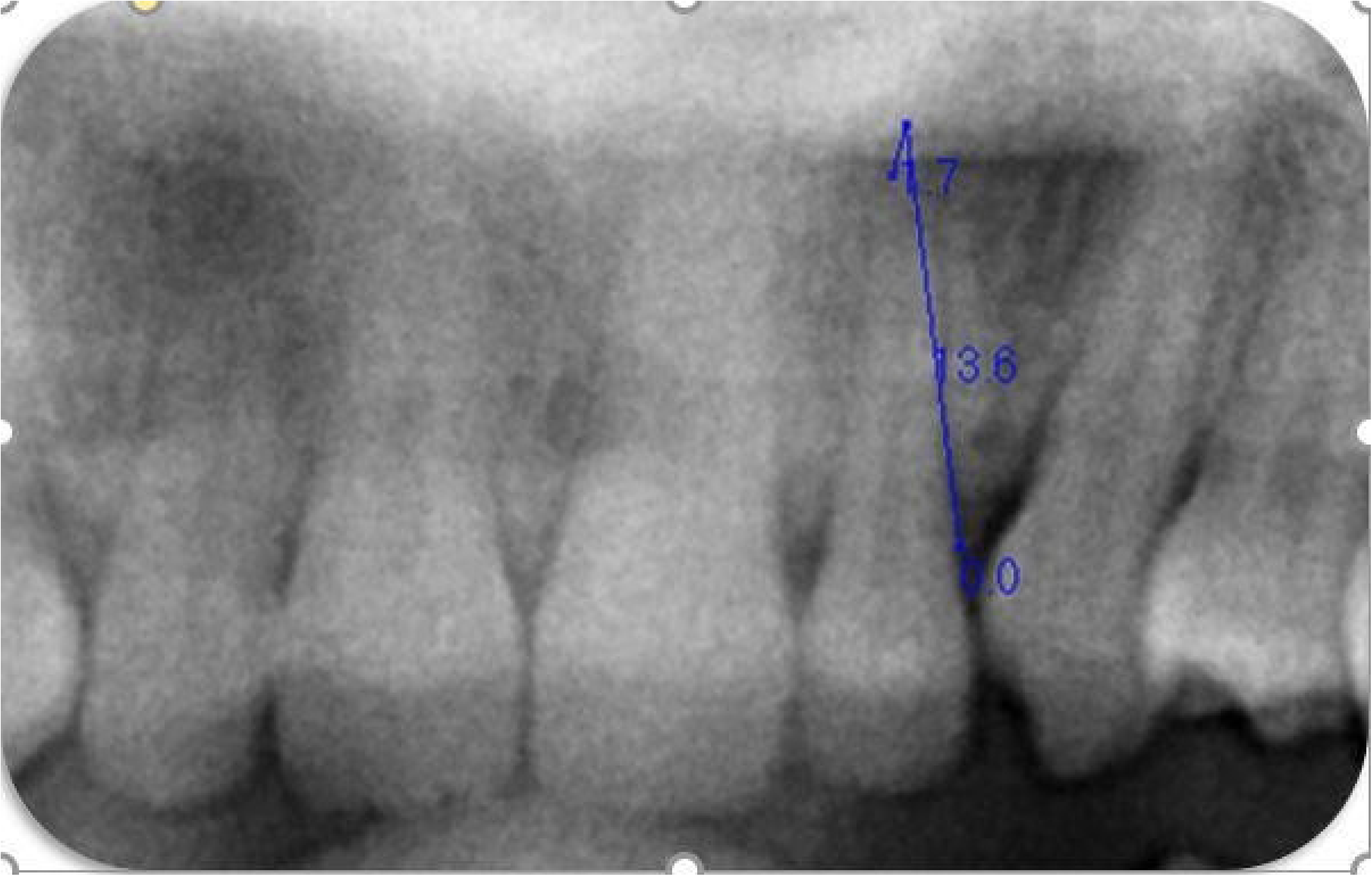
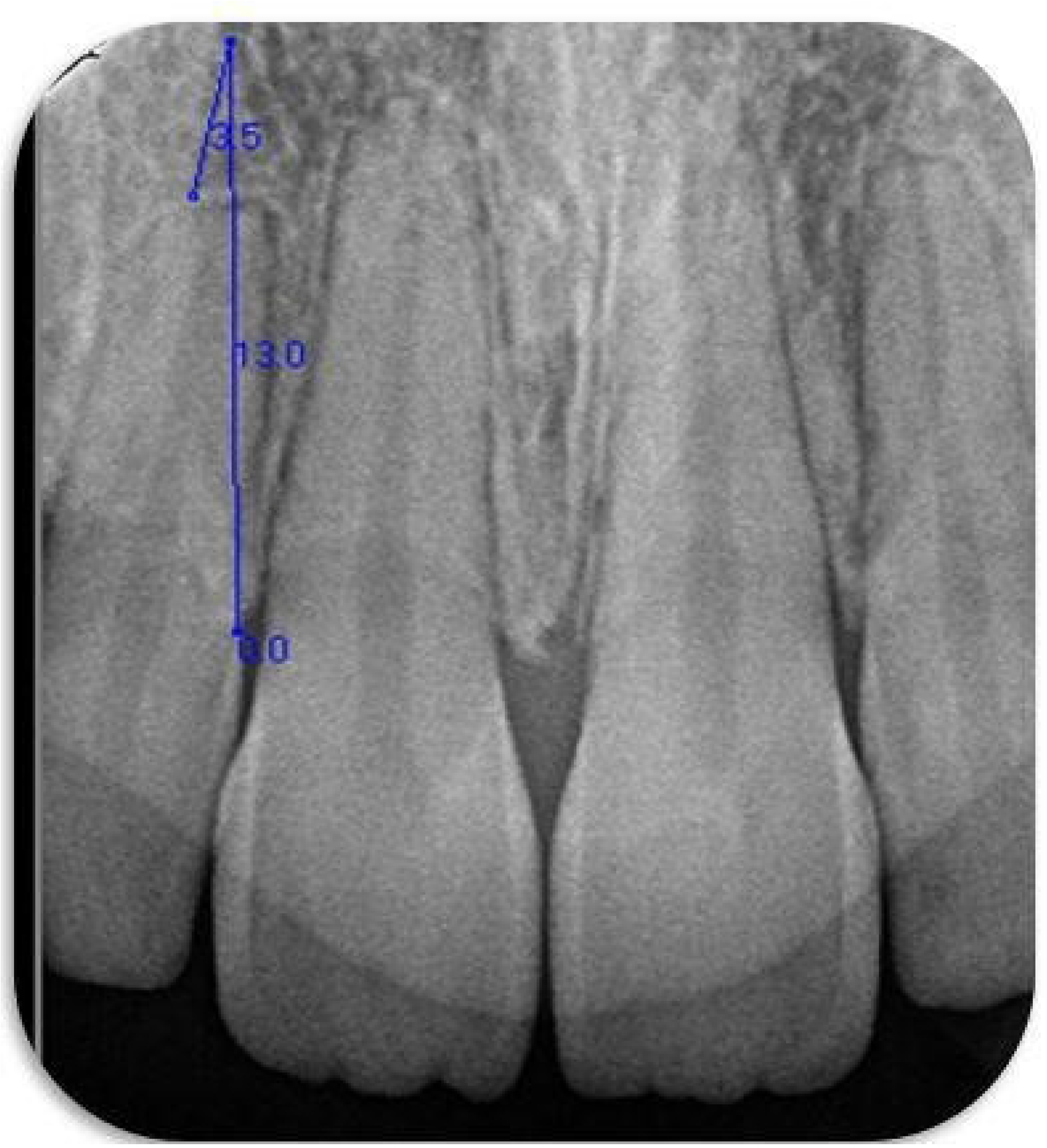
Two Periapical radiographs for two cases are representing the methodology of assessment of apical root resorption with comparing; A and B, show post-treatment radiographs of the same type.

### 2.5 Propensity Score Matching

Propensity score matching was used to improve comparability between groups and address potential biases. Among the matching variables were:

- Treatment type (fixed appliances vs. Invisalign).
- Patient demographics (age and gender).
- Malocclusion classification (Class I, II, or III).
- Extraction status, treatment duration, and frequency of orthodontic visits.

30 cases were maintained in each group post matching, ensuring comparability across important factors.

### 2.6 Data and Statistical Analysis

The data distribution’s normality was evaluated using the Shapiro-Wilk test. Parametric or non-parametric tests were used considering the significance of these findings:

- Continuous variables were compared using independent t-tests or Mann-Whitney U tests.
- Categorical variables were analyzed using chi-square tests.
- Statistical significance was set at p < 0.05.

## 3. Results

The analysis included 203 cases, of which 30 were treated with Invisalign and 173 with fixed appliances. The patients began their orthodontic treatment between January 2010 and December 2020. First, 116 cases with fixed appliances and 17 with Invisalign were disqualified because of radiographs that were missing, patient information that was not complete, age restrictions, ongoing treatments, impacted canines, the use of mini screws, unclear radiographs, or previous maxillofacial surgeries.

The gender distribution of the remaining 60 matched cases was 44% male and 56% female. Class I malocclusion was the most common condition (46%), followed by Class II (43%) and Class III (11%). Patients were between the ages of 15 and 50, with the largest age group being those in the 15–25 age range. External root resorption was noted in 83% of cases; only 17% showed no signs of resorption. Interestingly, Class II malocclusion showed 100% root resorption, followed by Class I (84%) and Class III (77%) (Figure 2).

**Figure 2.**
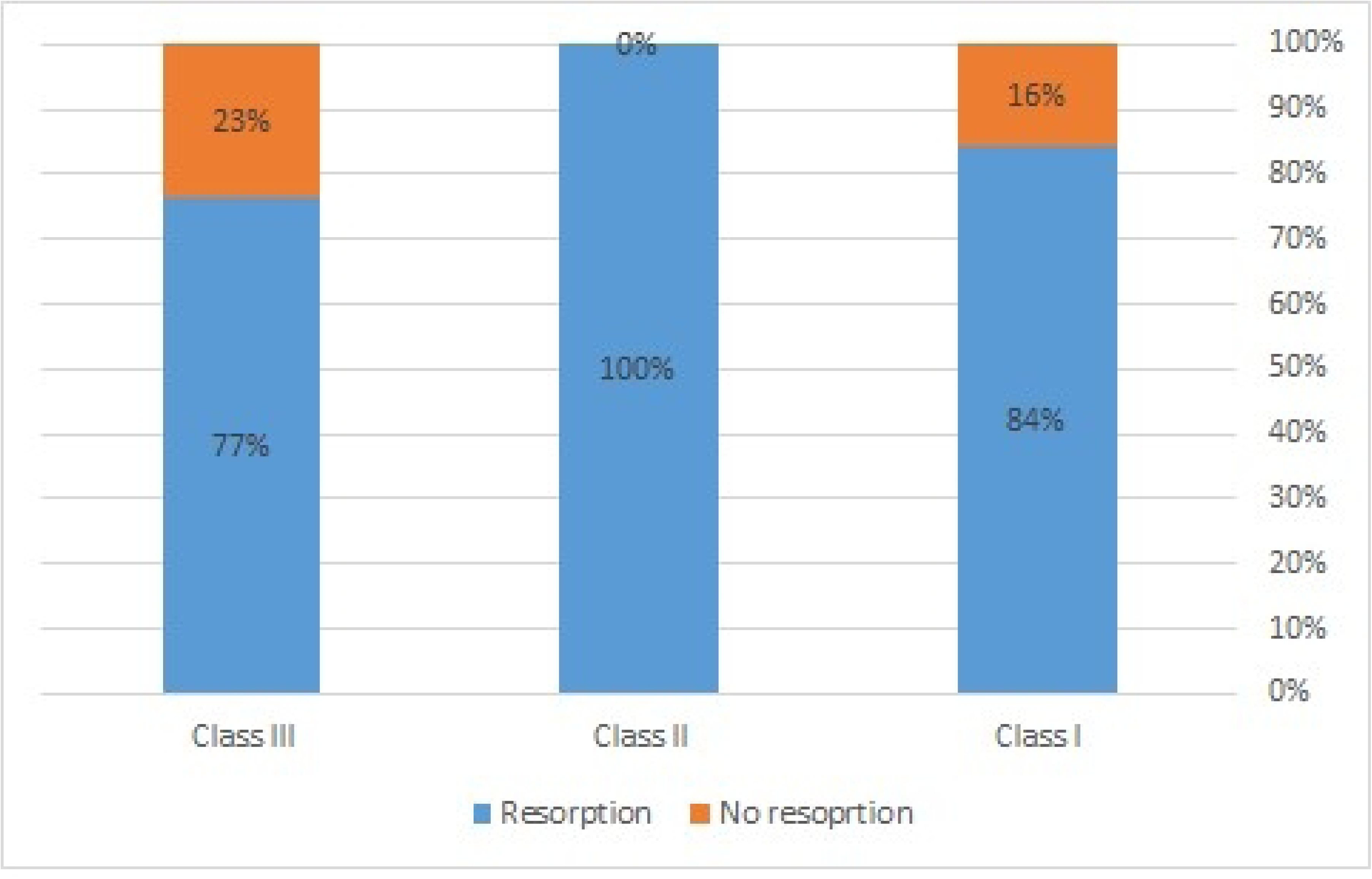
The bar chart represents the molar classification vs root resorption. class III have (77%) root resorption, class II with (100%) root resorption, class I (84%).

### 3.1 Comparison Between Treatment Modalities

Table 1 indicates that 28 Invisalign cases (92%) experienced external root resorption, while 23 fixed appliance cases (77%). The incidence of root resorption did not significantly differ between the two treatment groups, according to statistical analysis (p = 0.12). Invisalign cases showed shorter treatment durations (mean 18.2 ± 2.5 months) than fixed appliance cases (mean 22.6 ± 3.1 months; p = 0.045). Treatment durations varied across the study population. Specifically, 15.71% of treatments were completed within one year, 40% lasted two years, 35.71% extended to three years, and 8.57% took four years or more. Additionally, compared to the fixed appliance group (14.7 ± 2.0 visits; p = 0.038), the Invisalign group required significantly fewer orthodontic visits (10.3 ± 1.4 visits) (Figure 3).

**Figure 3.**
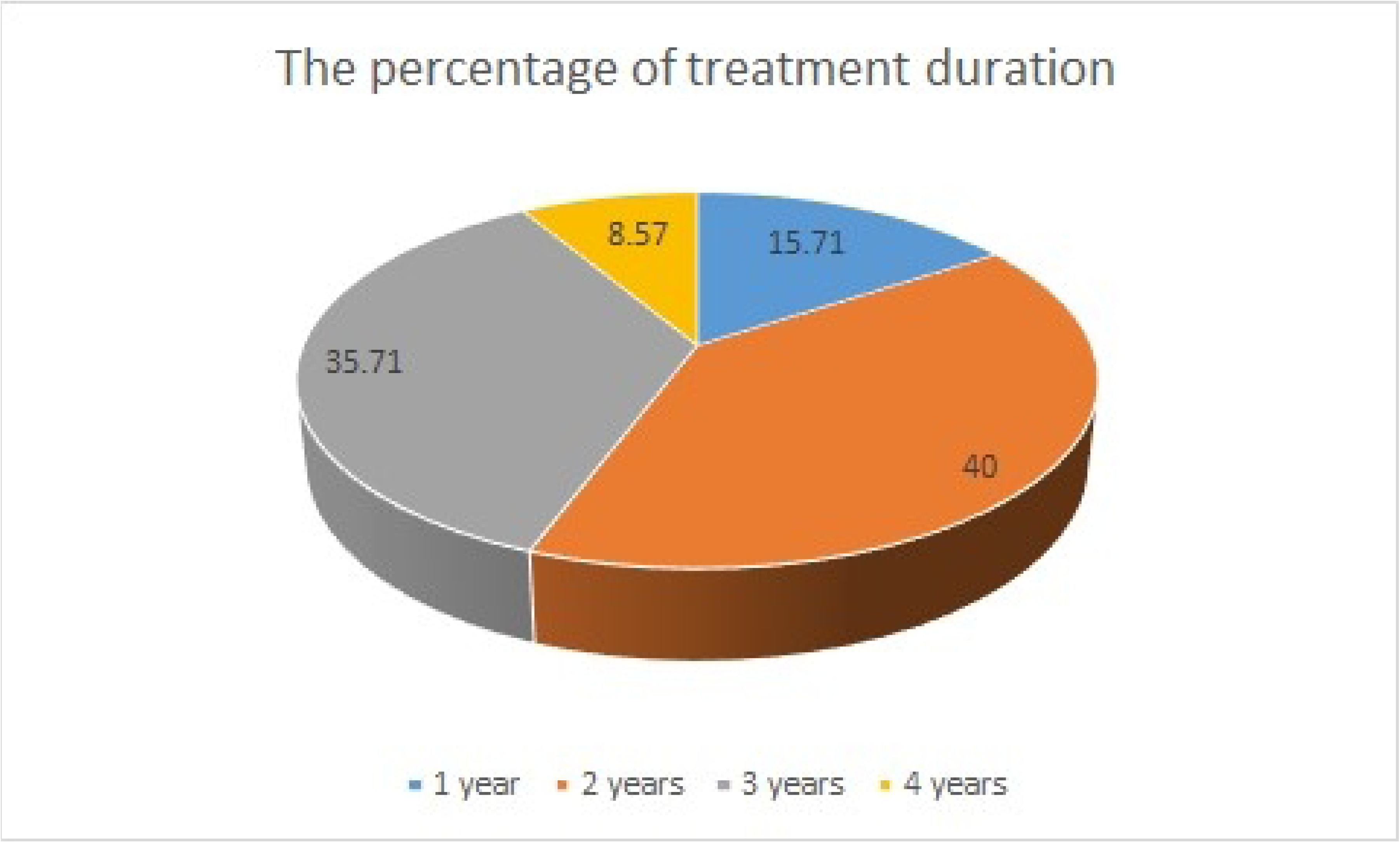
The pie chart represents the percentage of treatment duration of fixed appliances and clear aligner Invisalign. 1-year duration of treatment for fixed appliances and clear aligner Invisalign represent 15.71% .2 years’ duration of treatment for fixed appliances and clear aligner Invisalign represent 40%. 3 years’ duration of treatment for fixed appliances and clear aligner Invisalign represent 35.71. 4 years and above duration of treatment for fixed appliances and clear aligner Invisalign represent 8.57%.

**Table 1.** Comparison of Treatment Variables Between Invisalign and Fixed.

| Variable | Invisalign Group<br>(n=30) | Fixed Appliance<br>Group (n=30) | p-value |
| --- | --- | --- | --- |
| Root Resorption (%) | 92% (28 cases) | 77% (23 cases) | 0.12 |
| Treatment Duration<br>(Months) | $18.2 \pm 2.5$ | $22.6 \pm 3.1$ | 0.045 |
| Number of Visits | $10.3 \pm 1.4$ | $14.7 \pm 2.0$ | 0.038 |

### 3.2 Root Resorption and Extraction Status

Root resorption was seen in 26 (88%), as opposed to 23 (78%) of the patients who had their teeth extracted during treatment. However, the difference was not statistically significant (p = 0.14). These findings suggest that extractions may increase the likelihood of root resorption, but further data is needed for definitive conclusions (Figure 4).

**Figure 4.**
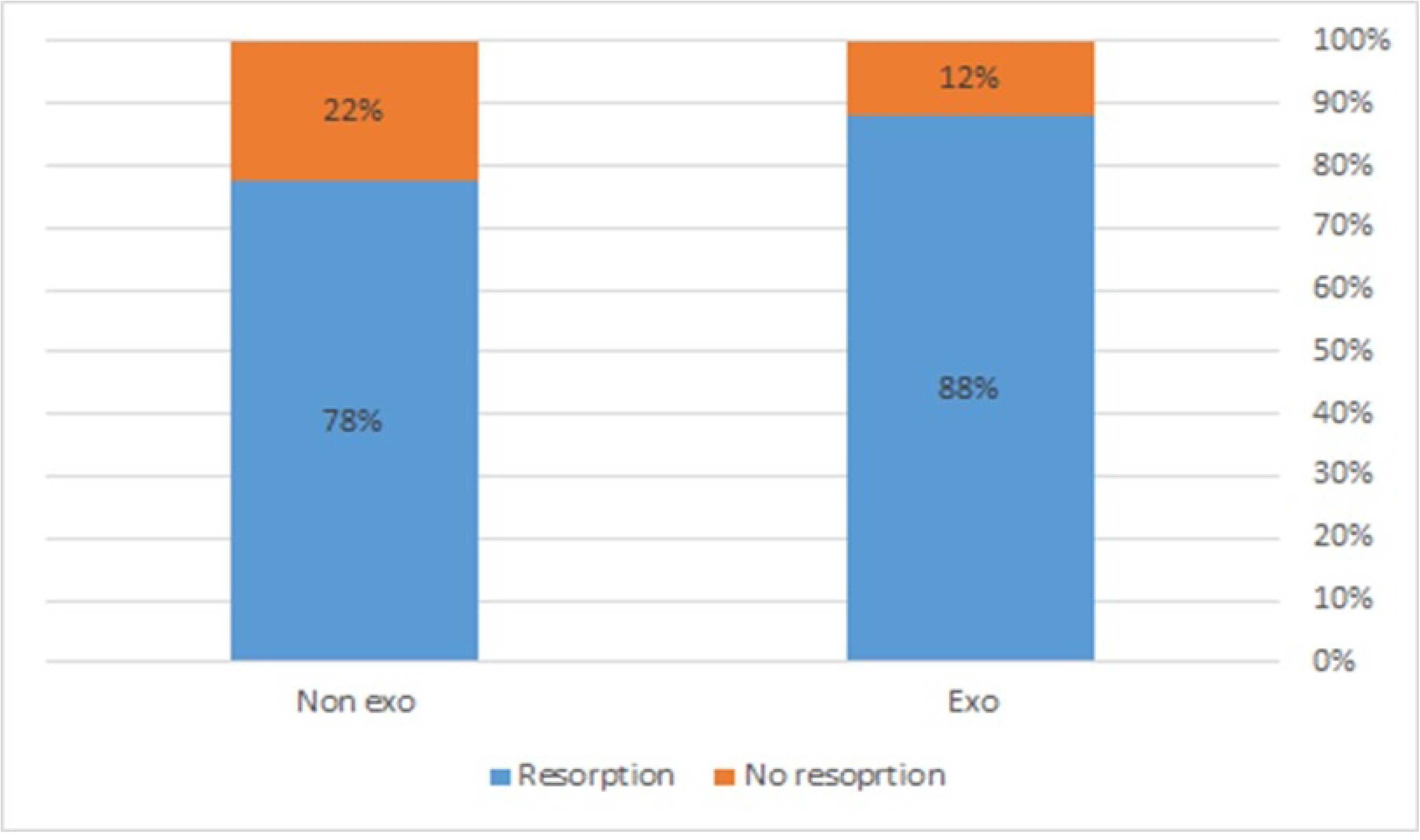
The root resorption of the cases has extracted teeth vs no extracted teeth. 88% cases have extracted teeth with root resorption while 12% no resorption. 78% cases have not extracted teeth with root resorption while 22% no resorption.

### 3.3 Gender Differences

The prevalence of root resorption was higher in females than in males, with 84.62% of cases occurring in female patients and 80.65% in males. Gender and the degree of root resorption did not significantly significant (p > 0.05).

### 3.4 Propensity-Matched Analysis

Using a propensity score-matching analysis, the 60 matched cases (30 in each group) were examined. Root resorption rates were higher in the Invisalign group (92%) compared to the fixed appliance group (77%) (Figure 5). However, this difference was not statistically significant (p = 0.12). The groups’ treatment durations and the number of teeth impacted were similar, as shown in Table 2.

**Figure 5.**
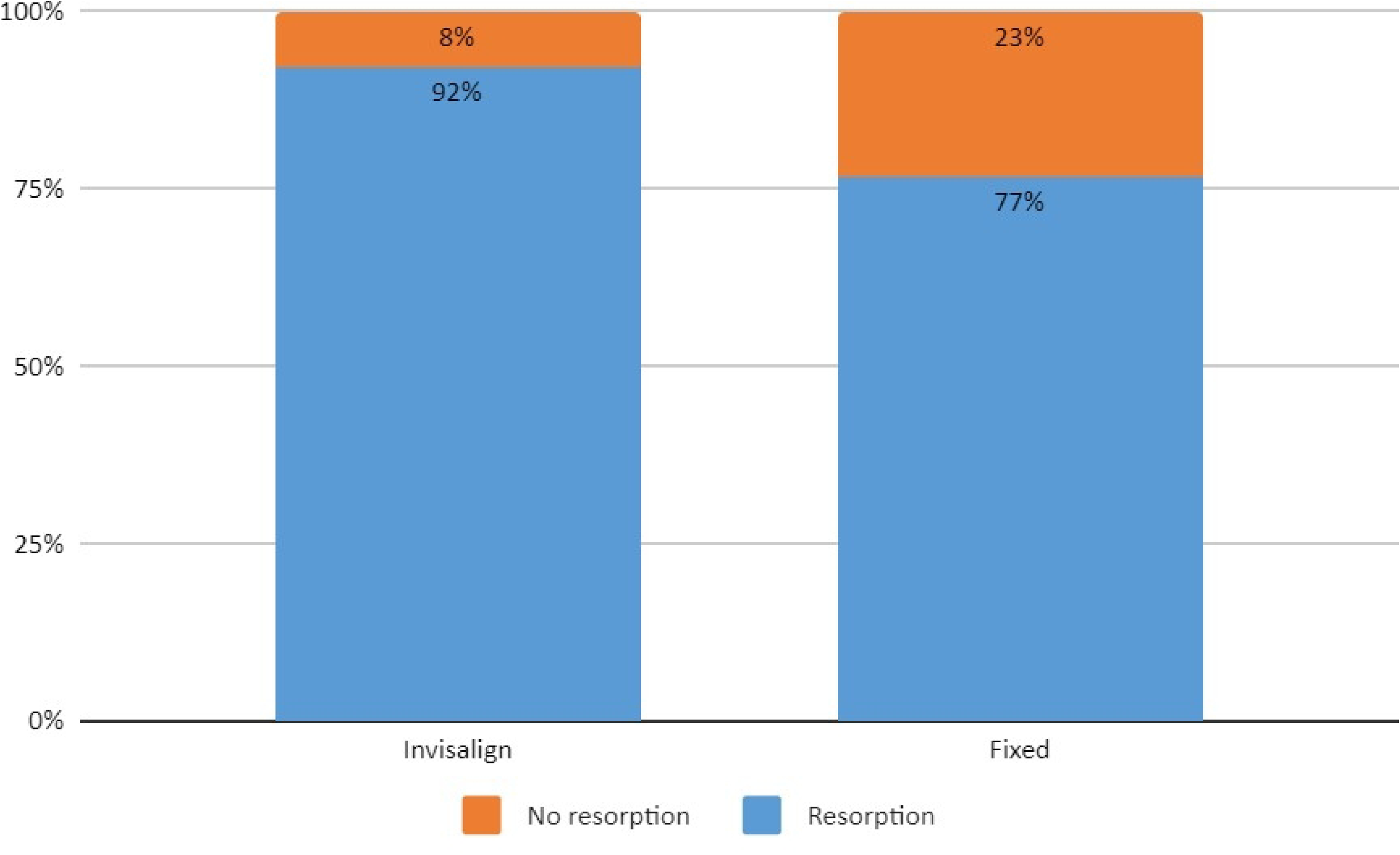
The bar chart represents matching cases .92% root resorption and 8% no root resorption of Invisalign cases vs 77% root resorption and 23% no root resorption of fixed appliances cases.

**Table 2.** Propensity-Matched Analysis of Root Resorption.

| Variable | Invisalign<br>Group (n=30) | Fixed Appliance<br>Group (n=30) | p-value |
| --- | --- | --- | --- |
| Root Resorption (%) | 92% (28<br>cases) | 77% (23 cases) | 0.12 |
| Maxillary Anterior Resorption<br>(%) | 88% (26<br>cases) | 75% (23 cases) | 0.15 |
| Number of Affected Teeth | 4.1 ± 1.0 | 5.6 ± 1.3 | 0.033 |

## 4. Discussion

Recently, there has been a lot of interest in the potential for root resorption when receiving orthodontic treatment with clear aligners like Invisalign. This problem has been the subject of numerous studies, which have shed light on its prevalence and seriousness. Loss of 1–2 mm of root length is considered clinically significant; severe cases (>4 mm) are uncommon but impact roughly 1–5% of patients [11].

A retrospective study using cone-beam computed tomography (CBCT) verified that apical root resorption can happen when clear aligners are used in orthodontic treatments [12]. Based on pre- and post-treatment radiographic evaluations, we found that external root resorption occurred in 92% of Invisalign cases and 77% of fixed appliance cases. These findings are consistent with earlier studies, like the one conducted by Flower et al., which showed that traditional orthodontic appliances were associated with a higher incidence of root destruction [13]. Similar to our findings, Krieger et al. (2014) reported that 54% of cases treated with clear aligners showed signs of root resorption [14].

Interestingly, other studies have shown divergent results. In contrast to fixed appliances (6.97 ± 3.67%), Yi et al. found lower levels of root resorption in Invisalign cases (5.13 ± 2.81%), which they attributed to the aligners’ sporadic forces [15].

Li et al. also used CBCT imaging to find significantly lower apical root resorption in clear aligner cases (56.3%) compared to fixed appliances (82.1%). Variations in study designs, imaging modalities, and the severity of malocclusions treated could be the cause of these disparities [16].

Maxillary incisors are especially vulnerable to resorption during orthodontic treatment because of their prominent position, frequent participation in movements related to extraction, and increased force sensitivity. Our findings confirmed that maxillary incisors were the most affected teeth, with minimal resorption observed in mandibular first molars. Previous studies on tooth-specific susceptibility during orthodontic treatments are consistent with this pattern [17].

The prediction and management of root resorption are complicated by its multifactorial nature. Significant contributions are made by elements like the length of treatment, the kind and extent of tooth movement, the status of extractions, and patient-specific traits (such as bone density and general health). The difficulty is in applying conservative forces, especially in fixed appliance treatments, but once orthodontic forces are removed, root resorption usually stops. There is evidence in the literature that the intermittent forces used by clear aligners, as opposed to the continuous forces used by fixed appliances, result in less resorption [18].

### 4.1 Strengths and limitations

Because of its retrospective design and reliance on panoramic radiographs, this study had inherent limitations even though the sample size was increased to increase statistical reliability. Despite its practicality, panoramic imaging is less precise than CBCT, which could lead to inconsistent measurements of root resorption. Furthermore, our capacity to examine important elements like cortical bone interaction and tooth movement type (e.g., tipping vs. bodily movement) was limited by the lack of cephalometric data. These limitations indicate the need for prospective studies using larger, multi-center samples and advanced imaging techniques to confirm and build upon these findings, without invalidating the findings.

## 5. Conclusion

Orthodontic treatment is essential for regaining oral health, function, and appearance. In contrast to patients treated with traditional fixed appliances, this study examined the frequency and severity of external root resorption in patients treated with Invisalign clear aligners. Invisalign cases had a higher frequency (92%) than fixed appliance cases, which showed more severe resorption in 77% of cases, but they also had shorter treatment durations and fewer orthodontic visits.

The results emphasize that when selecting treatment modalities, clinicians should take into account the risk factors specific to each patient, including the degree of malocclusion, the need for extraction, and the length of treatment. Even though this study’s findings offer insightful information, more research with larger, prospective cohorts, standardized imaging procedures, and comprehensive cephalometric analyses are essential to better understand the factors contributing to root resorption and to develop strategies for its prevention.

## Data Availability

Data cannot be shared publicly because of IRB rules in the instritution. Data are available from the radiographs Institutional Data Access / Ethics Committee (contact via email only) for researchers who meet the criteria for access to confidential data.

NA

## Acknowledgements

We acknowledge Dr. Ahmad and Dr. Mohammed Ibrahim for their clinical support and for providing Invisalign cases from the Jeddah population.

## Competing Interests

The authors have declared that no competing interests exist.

## Notes

### Competing Interest Statement

The authors have declared no competing interest.

### Clinical Trial

NA

### Clinical Protocols

NA

### Funding Statement

The author(s) received no specific funding for this work.

### Author Declarations

The Faculty of Dentistry's Research Ethics Committee at king Abdulaziz University provided its approval to the research being performed (Approval number: 161-12-20).

